# Contribution of genetic and environmental factors to the onset of preclinical Alzheimer’s disease - a monozygotic twin study

**DOI:** 10.1101/2020.06.12.20129346

**Authors:** Elles Konijnenberg, Jori Tomassen, Anouk den Braber, Mara ten Kate, Maqsood M. Yaqub, Sandra D. Mulder, Michel G. Nivard, Hugo Vanderstichele, Adriaan A. Lammertsma, Charlotte E. Teunissen, Bart N.M. van Berckel, Dorret I. Boomsma, Philip Scheltens, Betty M. Tijms, Pieter Jelle Visser

## Abstract

**Objective:** To study the genetic contribution to the start of Alzheimer’s disease as signified by abnormalities in amyloid and tau biomarkers in cognitively intact older identical twins.

**Methods:** We studied in 96 monozygotic twin-pairs relationships between Aβ aggregation as measured by the ratio Aβ1-42/1-40 in cerebrospinal fluid (CSF) and positron emission tomography (PET), and CSF markers for Aβ production (BACE1, Aβ1-40 and 1-38) and tau. Associations amongst markers were tested with Generalized Estimating Equations including a random effect for twin status, adjusted for age, gender, and APOE ε4 genotype. We used twin analyses to determine relative contributions of genetic and/or environmental factors to AD pathophysiological processes.

**Results:** Twenty-seven individuals (14%) had an abnormal amyloid-PET, and 14 twin-pairs (15%) showed discordant amyloid status. Within twin-pairs, Aβ production markers and total-tau (t-tau) levels strongly correlated (r range 0.76, 0.88; all p<0.0001), and Aβ aggregation markers and 181-phosphorylated-tau (p-tau) levels correlated moderately strong (r range 0.49, 0.52; all p<0.0001). Cross-twin cross-trait analysis showed that Aβ1-38 in one twin correlated with Aβ1-42/1-40 ratios, t-tau and p-tau levels in their co-twins (r range 0.18, 0.58; all p<.07). Within-pair differences in Aβ production markers related to differences in tau levels (r range 0.49, 0.61; all p<0.0001). Twin discordance analyses suggest that Aβ production and tau levels show coordinated increases in very early AD.

**Interpretation:** Our results suggest a substantial genetic/shared environmental background contributes to both Aβ and tau increases, suggesting that modulation of environmental risk factors may aid in delaying the onset of AD pathophysiological processes.

## 1. Introduction

Aggregation of amyloid-beta 1-42 (Aβ1-42) in the brain can start up to 20 years before the onset of dementia^1^, followed by tau pathology and cognitive decline^2,3^. The mechanisms leading to Aβ aggregation and tau pathology are not fully understood, while such knowledge is crucial for the development of primary and secondary prevention strategies. Therefore it is important to understand the contribution of genetic and environmental factors in the early development of the disease, when biomarkers for Aβ and tau are changing and cognition is still intact. Twin studies suggested that genetic factors may explain 80% of the variance in AD-type dementia^4,5^, but it remains largely unknown whether this is also the case for AD biomarkers in elderly cognitively normal individuals^6,7^.

In autosomal dominant variants of Alzheimer’s disease (ADAD), Aβ aggregation is associated with increased Aβ production. In sporadic Alzheimer’s disease (AD), however, impaired clearance is suggested to drive the disease^8,9^. Nevertheless, iPSC models in sporadic AD suggest that increased Aβ production and tau secretion are also involved^10^. In people, higher cerebrospinal fluid (CSF) BACE1 levels, the rate-limiting enzyme in Aβ production, have been associated with higher CSF concentrations of Aβ peptides^11^, and subsequent aggregation of Aβ1-42 in individuals with initially normal Aβ CSF levels^12^.

While it is generally assumed that tau follows Aβ pathology^13^, CSF levels of total-tau (t-tau) and 181-phosphorylated-tau (p-tau) are also increased early in the disease^14^. Possibly, such early increases in CSF tau and Aβ levels are driven by common upstream pathophysiological processes^15^, and in order to identify such upstream processes it is important to determine the relative contributions of genetic and environmental factors to early changes in these AD pathophysiological processes. A monozygotic twin design provides a unique approach to study the role of genetic and environmental factors in disease development. Since monozygotic twins are genetically identical, differences for a trait within twin-pairs result from differences in unique environmental exposure.

Our objective was to investigate associations between Aβ production, Aβ aggregation and CSF tau pathology in cognitively intact older monozygotic twins, to determine the role of genetic and environmental factors on these associations. We used twin discordance models to test the dynamic relationship between Aβ production, Aβ aggregation markers: because twin concordance for AD increases with age^4^, we hypothesized that if Aβ production precedes Aβ aggregation, then in twin-pairs discordant for Aβ aggregation, the discordant twin with normal Aβ levels would already show signs of increased Aβ production markers compared to twin-pairs with normal Aβ markers. Furthermore, if Aβ and tau pathology are driven by common upstream pathophysiological processes, then higher levels of Aβ markers in one twin should be related to higher levels of tau levels in the co-twin in cross-twin cross-trait (CTCT) analysis. If associations between tau and Aβ markers are driven by unique environmental factors, then twin differences in these markers should be correlated. Furthermore, we investigated whether these analyses depend on the modality used to determine Aβ abnormality (i.e., positron emission tomography (PET) or CSF).^16^

## 2. Methods

### 2.1. Participants

Monozygotic twins were invited from the Netherlands Twin Register (NTR)^17^ to participate in the PreclinAD study as part of the Innovative Medicine Initiative (IMI) European Information Framework for AD (EMIF-AD) project (http://www.emif.eu/)^18^. Inclusion criteria were age 60 years and older, a delayed recall score of > -1.5 SD of demographically adjusted normative data of the Consortium to Establish a Registry for Alzheimer’s Disease (CERAD) 10 word list^19^, a Telephone Interview for Cognitive Status modified (TICS-m) score of 23 or higher^20^, a 15-item Geriatric Depression Scale (GDS) score of <11^21^, and a Clinical Dementia Rating (CDR) scale of 0 with a score on the memory sub domain of 0^22^. Exclusion criteria were any physical, neurological or psychiatric condition that could lead to interference with normal cognition in aging. Monozygotic twins were asked to collect buccal cell samples for DNA extraction to confirm zygosity. For the present study we included all individuals who had an amyloid measurement available (n=197).

### 2.2. Ethical considerations

Informed consent was obtained from all participants. The study was approved by the Central Ethics Committee on Research Involving Human Subjects of the VU University Medical Centre, Amsterdam, an Institutional Review Board certified by the U.S. Office of Human Research Protections (IRB number IRB00002991 under Federal-wide Assurance-FWA00017598). The research was performed according to the principles of the Declaration of Helsinki and in accordance with the Medical Research Involving Human Subjects Act and codes on ‘good use’ of clinical data and biological samples as developed by the Dutch Federation of Medical Scientific Societies. The study was registered in the EU Clinical Trials Register (EudraCT) with number 2014-000219-15.

### 2.3. Cerebrospinal fluid analysis

To determine Aβ aggregation in CSF we used the Aβ 1-42/1-40 ratio with lower values indicating abnormality^23^. As markers for Aβ production we used BACE1 concentrations in CSF, as well as concentrations of Aβ 1-40 (Aβ40) and 1-38 (Aβ38)^24^. For tau pathology we used CSF t-tau and p-tau^25,26^, which reflect different aspects of tau pathology^25,27-29^, with especially higher values for p-tau being indicative for tau aggregation. CSF samples were collected in 126 (62%) participants through a lumber puncture, performed between 10am and 2pm, after at least two hours of fasting. Maximal 20 ml CSF was collected in Sarstedt polypropylene syringes using a Spinocan 25 Gauge needle in one of the intervertebral spaces between L3 and S1. Samples were centrifuged at 1300-2000 g at 4**°**C for 10 minutes and supernatants were then stored in aliquots at -80**°**C until analysis^30^. A maximum of 2 hours was allowed between lumbar puncture and freezing. Levels of Aβ 1-38, 1-40, 1-42, BACE1, t-tau and p-tau were analyzed using commercial kits from Euroimmun AG (Lübeck, Germany) according to manufacturer instructions^31^. All samples were measured in kits from the same lot.

### 2.4. [^18^F]flutemetamol positron emission tomography

To determine Aβ aggregation on PET imaging we used parametric non-displaceable binding potential (BP_ND_) of the [^18^F]flutemetamol tracer, with higher binding of Aβ radioligands indicating the presence of plaques^32-34^. 196 subjects (94 twin-pairs) had [^18^F]flutemetamol PET available. In general, PET scanning was performed on the same day as the lumbar puncture, except for 26 subjects, due to technical issues (range 2.2 months before to 6.7 months after lumbar puncture). PET scans were acquired using a Philips Ingenuity TF PET-MRI camera (Philips Healthcare, Cleveland, USA). All subjects were scanned under standard resting conditions (eyes closed in dimmed ambient light) from 0 to 30 minutes and from 90 to 110 minutes after intravenous injection of 185 MBq (±10%) [^18^F]flutemetamol^34^. After data acquisition, the first emission scan was reconstructed into 18 frames of increasing length (6×5, 3×10, 4×60, 2×150, 2×300 and 1×600 s.) using the standard LOR-RAMLA reconstruction algorithm for the brain. Using the same reconstruction algorithm, the second scan was reconstructed into 4 frames of 5 minutes each. Subsequently, data from the two scans were combined into a single image data set after co-registration using Vinci Software 2.56 (Max Planck Institute for Neurological Research, Cologne, Germany). Parametric non-displaceable binding potential (BP_ND_) images were generated from the entire image set using receptor parametric mapping^35,36^. Cerebellar grey matter, defined on a T1-weighted structural MRI scan obtained immediately prior to the PET scan, was used for attenuation correction of the PET data and as reference tissue^37^. T1-based VOIs using the Hammers atlas implemented in PVElab software were projected onto the [^18^F]flutemetamol parametric images to extract regional values^38^. A global BP_ND_ was calculated based on the volume-weighted average of frontal (i.e. superior, middle, and inferior frontal gyrus), parietal (i.e., posterior cingulate, superior parietal gyrus, postcentral gyrus, and inferolateral remainder of parietal lobe), and temporal (i.e. parahippocampal gyrus, hippocampus, medial temporal lobe, superior, middle, and inferior temporal gyrus) cortical regions^39^. We classified twins as amyloid positive (abnormal) or negative (normal) by visual read of the [^18^F]flutemetamol scans. Rating was performed on the parametric BP_ND_ images by three readers (nuclear physician or radiologist) all trained according to GEHC guidelines^40,41^. For 15 cases not all readers agreed, here we used the consensus rating of 2 readers. In quantitative analyses we used the global BP_ND_.

### 2.6. APOE genotyping

To assess APOE ε4 allele carriership, the major genetic risk factor for sporadic AD, all subjects were genotyped on the Affymetrix Axiom array and the Affymetrix 6 array^42^. These were first cross chip imputed following the protocols as described by Fedko and colleagues^43^ and then imputed to HRC with the Michigan Imputation server^44^. APOE genotype was assessed using imputed dosages of the SNP rs429358 (APOE ε4, imputation quality = 0.956) and rs7412 (APOE ε2, imputation quality = 0.729)^45^.

### 2.7. Statistical analysis

We used Generalized Estimating Equations (GEE) to test amyloid-PET group differences for demographic variables, including random effect for twin status. We then assessed associations between Aβ and tau markers across the total group, using GEE including random effect for twin status. Analyses were performed unadjusted (model 1) and adjusted for age, gender, and APOE ε4 genotype (model 2) when applicable^46^. Next we performed three types of twin analyses. Monozygotic twin-pair correlations (i.e. correlations for a trait between twin 1 and twin 2 across the group) for CSF (n=54 pairs) and PET amyloid (n=94 pairs) were assessed using Pearson’s correlations, which provides a proxy of variance explained in a trait by the combination of shared genetic and shared environmental factors. The correlation coefficient-1 indicates the percentage of the trait explained by unique environmental factors. Partial correlations were also calculated adjusting for age, gender and APOE ε4 genotype. PET data and CSF tau data were log-transformed to improve normal distribution of the data. When two variables showed a significant association, we performed CTCT analysis to test whether levels of marker A in one twin could predict levels of marker B in the co-twin using OpenMx^47^. We also performed a monozygotic within-pair difference analysis^48^ when there was a significant association between variables. This analysis allows examining whether within-pair differences in marker A can be explained by within-pair differences in marker B. It provides insight into the contribution of unique environmental factors unique to a twin on the observed relation between traits^48^. Statistical analyses were performed in SPSS version 23 for Windows and R version 3.3.1, http://www.r-project.org/.

## 3. Results

### Sample description

Twenty-seven individuals (14%) had an abnormal visual read of the amyloid-PET scan. These subjects were older and had lower CSF Aβ1-42/1-40 ratios and higher p-tau levels compared to individuals with a normal PET scan (Table 1).

**Table 1.**
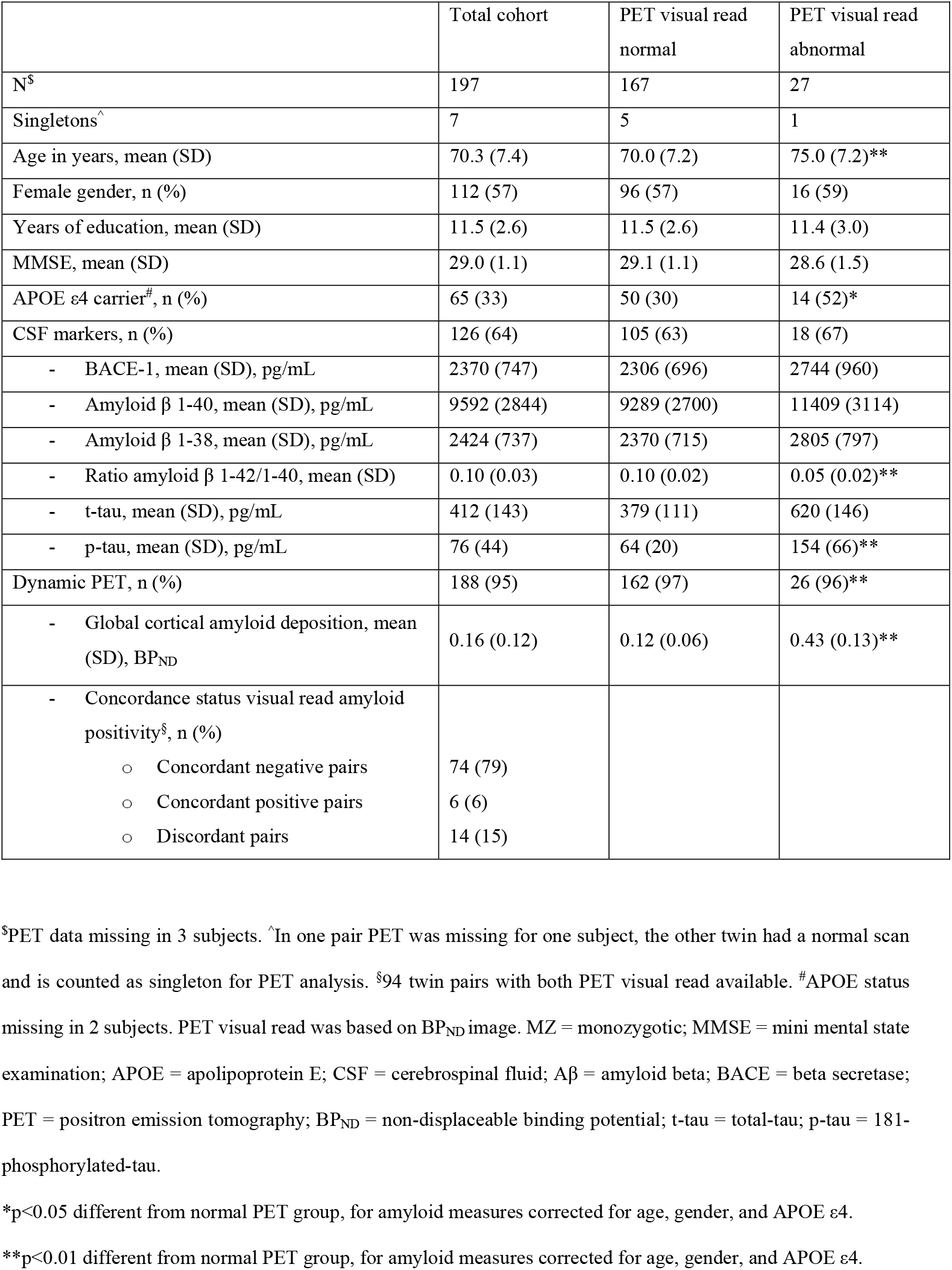
Cohort characteristics.

### Twin-pair correlations

Monozygotic twin-pair correlations (i.e. correlation for a trait across paired twins) were strong for Aβ production markers (r ranging between 0.76 and 0.85) and t-tau (r=0.70), and moderately strong for Aβ aggregation markers (r range 0.50, 0.52) and p-tau (r=0.49) (Fig. 2, Table 3). None of the markers tested were correlated across unrelated individuals (r range 0.05, 0.3; all p<0.99), and effect sizes remained similar when correcting for age, gender and APOE ε4 (Table 3).

**Table 2.**
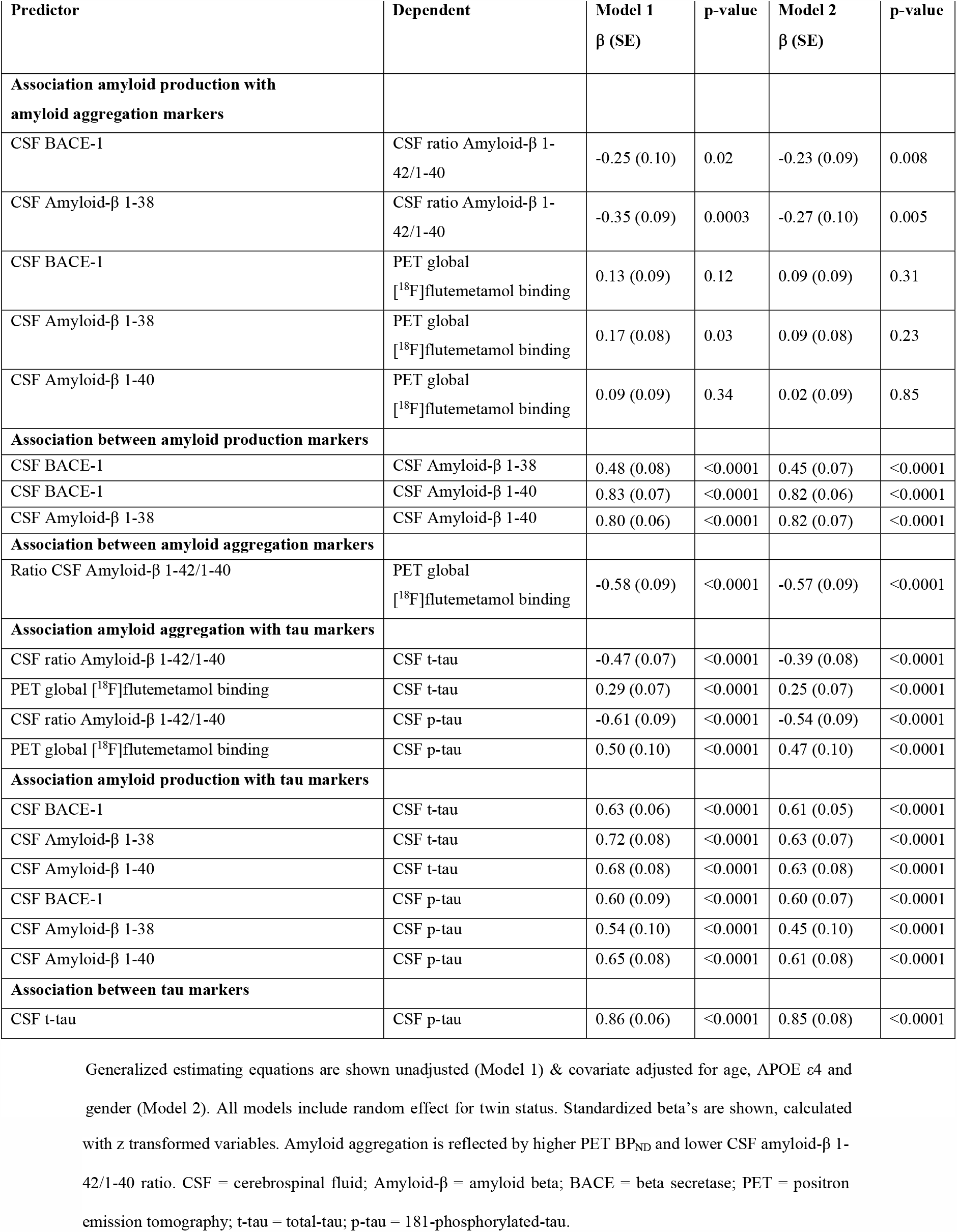
Association between amyloid production, amyloid aggregation and tau, among amyloid production, amyloid aggregation and tau in total cohort.

| Predictor | Dependent | Model 1<br>$\beta$ (SE) | p-value | Model 2<br>$\beta$ (SE) | p-value |
| --- | --- | --- | --- | --- | --- |
| **Association amyloid production with amyloid aggregation markers** |  |  |  |  |  |
| CSF BACE-1 | CSF ratio Amyloid-β 1-42/1-40 | -0.25 (0.10) | 0.02 | -0.23 (0.09) | 0.008 |
| CSF Amyloid-β 1-38 | CSF ratio Amyloid-β 1-42/1-40 | -0.35 (0.09) | 0.0003 | -0.27 (0.10) | 0.005 |
| CSF BACE-1 | PET global [$^{18}\text{F}$]flutemetamol binding | 0.13 (0.09) | 0.12 | 0.09 (0.09) | 0.31 |
| CSF Amyloid-β 1-38 | PET global [$^{18}\text{F}$]flutemetamol binding | 0.17 (0.08) | 0.03 | 0.09 (0.08) | 0.23 |
| CSF Amyloid-β 1-40 | PET global [$^{18}\text{F}$]flutemetamol binding | 0.09 (0.09) | 0.34 | 0.02 (0.09) | 0.85 |
| **Association between amyloid production markers** |  |  |  |  |  |
| CSF BACE-1 | CSF Amyloid-β 1-38 | 0.48 (0.08) | <0.0001 | 0.45 (0.07) | <0.0001 |
| CSF BACE-1 | CSF Amyloid-β 1-40 | 0.83 (0.07) | <0.0001 | 0.82 (0.06) | <0.0001 |
| CSF Amyloid-β 1-38 | CSF Amyloid-β 1-40 | 0.80 (0.06) | <0.0001 | 0.82 (0.07) | <0.0001 |
| **Association between amyloid aggregation markers** |  |  |  |  |  |
| Ratio CSF Amyloid-β 1-42/1-40 | PET global [$^{18}\text{F}$]flutemetamol binding | -0.58 (0.09) | <0.0001 | -0.57 (0.09) | <0.0001 |
| **Association amyloid aggregation with tau markers** |  |  |  |  |  |
| CSF ratio Amyloid-β 1-42/1-40 | CSF t-tau | -0.47 (0.07) | <0.0001 | -0.39 (0.08) | <0.0001 |
| PET global [$^{18}\text{F}$]flutemetamol binding | CSF t-tau | 0.29 (0.07) | <0.0001 | 0.25 (0.07) | <0.0001 |
| CSF ratio Amyloid-β 1-42/1-40 | CSF p-tau | -0.61 (0.09) | <0.0001 | -0.54 (0.09) | <0.0001 |
| PET global [$^{18}\text{F}$]flutemetamol binding | CSF p-tau | 0.50 (0.10) | <0.0001 | 0.47 (0.10) | <0.0001 |
| **Association amyloid production with tau markers** |  |  |  |  |  |
| CSF BACE-1 | CSF t-tau | 0.63 (0.06) | <0.0001 | 0.61 (0.05) | <0.0001 |
| CSF Amyloid-β 1-38 | CSF t-tau | 0.72 (0.08) | <0.0001 | 0.63 (0.07) | <0.0001 |
| CSF Amyloid-β 1-40 | CSF t-tau | 0.68 (0.08) | <0.0001 | 0.63 (0.08) | <0.0001 |
| CSF BACE-1 | CSF p-tau | 0.60 (0.09) | <0.0001 | 0.60 (0.07) | <0.0001 |
| CSF Amyloid-β 1-38 | CSF p-tau | 0.54 (0.10) | <0.0001 | 0.45 (0.10) | <0.0001 |
| CSF Amyloid-β 1-40 | CSF p-tau | 0.65 (0.08) | <0.0001 | 0.61 (0.08) | <0.0001 |
| **Association between tau markers** |  |  |  |  |  |
| CSF t-tau | CSF p-tau | 0.86 (0.06) | <0.0001 | 0.85 (0.08) | <0.0001 |

Generalized estimating equations are shown unadjusted (Model 1) & covariate adjusted for age, APOE ε4 and gender (Model 2). All models include random effect for twin status. Standardized beta's are shown, calculated
with z transformed variables. Amyloid aggregation is reflected by higher PET BP<sub>ND</sub> and lower CSF amyloid-β 1-42/1-40 ratio. CSF = cerebrospinal fluid; Amyloid- $\beta$ = amyloid beta; BACE = beta secretase; PET = positron emission tomography; t-tau = total-tau; p-tau = 181-phosphorylated-tau.

**Table 3.** Monozygotic twin-pair correlations.

| Markers | Correlation (95% CI)<br>Model 1 | Correlation (95% CI)<br>Model 2 |
| --- | --- | --- |
| BACE-1 | 0.76 (0.76 - 0.80) | 0.76 (0.75 - 0.79) |
| Amyloid-β 1-38 | 0.88 (0.88 - 0.89) | 0.85 (0.85 - 0.87) |
| Amyloid-β 1-40 | 0.78 (0.77 - 0.80) | 0.76 (0.76 - 0.78) |
| CSF ratio Amyloid-β 1-42/1-40 | 0.64 (0.64 - 0.67) | 0.50 (0.49 - 0.54) |
| PET global [<sup>18</sup>F]flutemetamol binding | 0.44 (0.44 - 0.50) | 0.52 (0.51 - 0.54) |
| CSF t-tau | 0.78 (0.78 - 0.81) | 0.70 (0.69 - 0.73) |
| CSF p-tau | 0.61 (0.59 - 0.69) | 0.49 (0.47 - 0.56) |

Correlation values for association of amyloid markers between one twin and its co-twin (Model 1), corrected for gender, age and APOE status (Model 2); all $p < 0.0001$ . Correlations are shown in figure 2. CSF = cerebrospinal fluid; PET = positron emission tomography; BACE = beta secretase; t-tau = total tau; p-tau = 181-phosphorylated-tau.

**Fig. 1.**
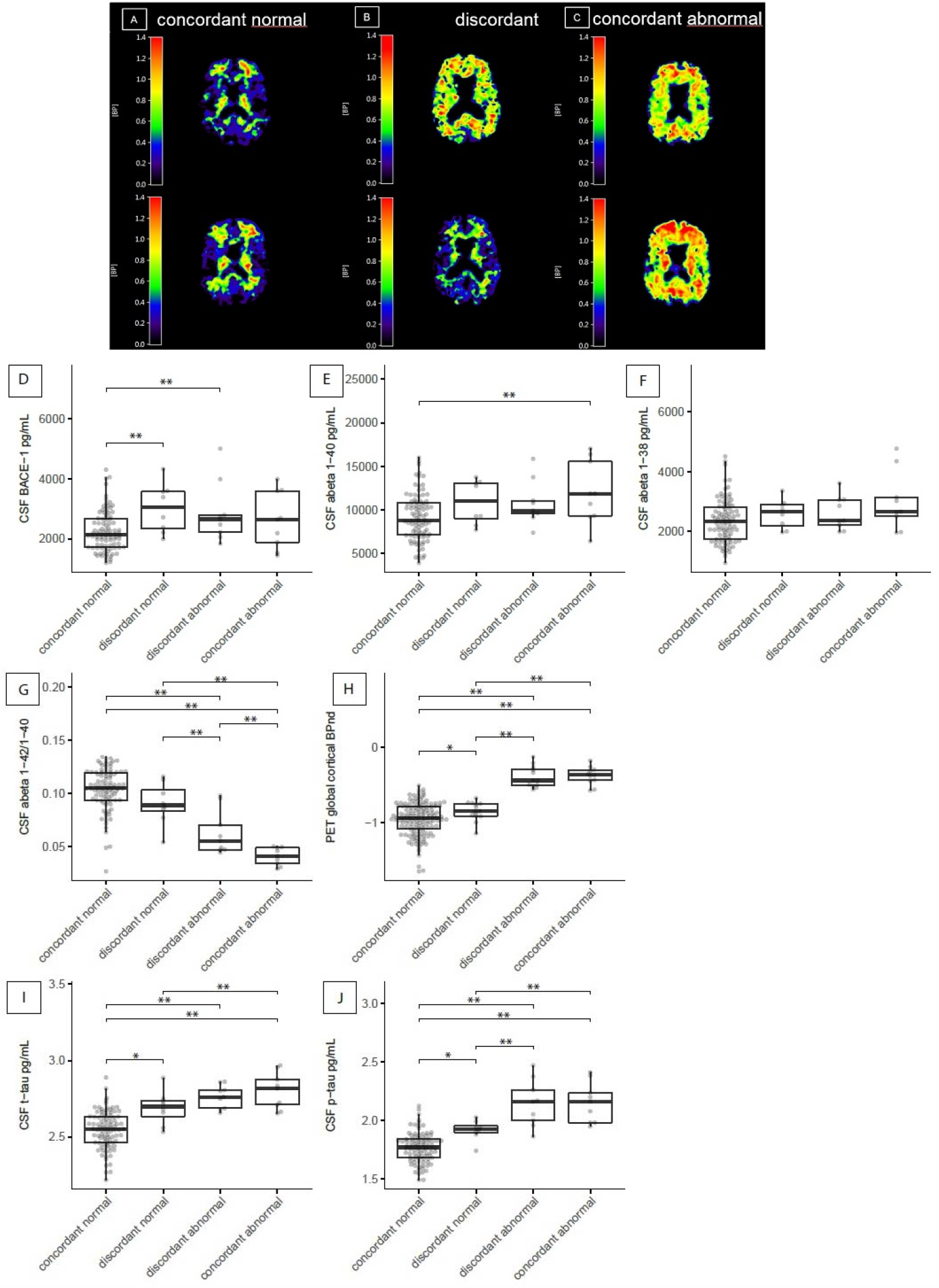
Patterns of amyloid production, amyloid aggregation and tau for twin discordance. [18F]flutemetamol PET images from a concordant twin pair (A) with a normal scan, a discordant pair (B) and a concordant pair with an abnormal scan (C). Boxplots show BACE1 (D); amyloid-β 1-40 (E); amyloid-β 1-38 (F); amyloid-β 1-42/1-40 ratio (G); global cortical PET binding (H); t-tau (I); and p-tau (J) for twins who have both a normal amyloid-PET scan (concordant normal, n=148 of which 93 have CSF markers), twins from a discordant pair with a normal amyloid-PET (discordant normal, n=14 of which 8 have CSF markers), twins from a discordant pair with abnormal amyloid-PET (discordant abnormal, n=14 of which 9 have CSF markers), and twin pairs who both have an abnormal amyloid-PET scan (concordant abnormal, n=12, of which 9 have CSF markers). All analyses for group comparisons were corrected for age, APOE ε4 and gender. *p<0.05; **p<0.01. CSF = cerebrospinal fluid; PET = positron emission tomography; BACE = beta secretase; t-tau = total-tau; p-tau = 181-phosphorylated-tau.

**Fig. 2.**
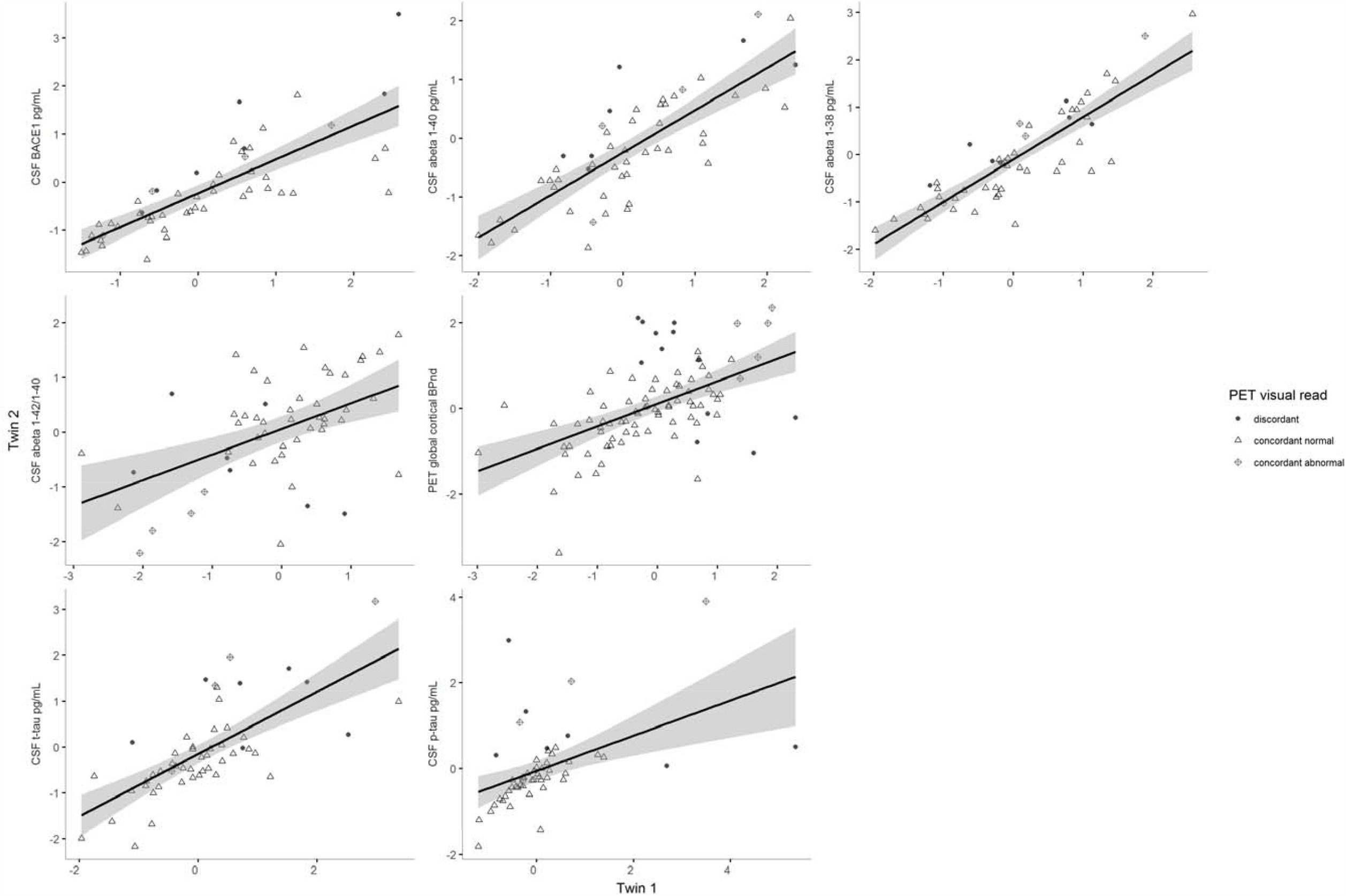
Monozygotic twin-pair correlations. Partial correlation values for association of amyloid markers between one twin and its co-twin, corrected for gender, age and APOE ε4 (Model 2); all p<0.0001. Each dot represents one twin pair. From left to right: CSF BACE1; CSF amyloid-β 1-40; CSF amyloid-β 1-38; CSF amyloid-β 1-42/1-40 ratio; global cortical PET binding (BP_ND_); CSF t-tau; CSF p-tau. CSF = cerebrospinal fluid; PET = positron emission tomography; BACE = beta secretase; t-tau = total-tau; p-tau = 181-phosphorylated-tau.

### Association between Aβ and tau markers across the total group

Across all subjects, higher levels of CSF BACE1 and Aβ38 were associated with lower CSF Aβ 1-42/1-40 ratios but not with amyloid-PET BP_ND_, and with higher levels of CSF t-tau and p-tau. Furthermore, lower CSF Aβ 1-42/1-40 ratios and higher amyloid-PET BP_ND_ were associated with higher levels of t-tau and p-tau, with p-tau showing a higher association with amyloid-PET BP_ND_ than t-tau (0.47 vs 0.25), Table 2).

### Discordant twin-pair analyses

Seventy-four pairs (79% of the 94 complete PET imaging pairs) had both a normal amyloid-PET visual read (i.e., concordant normal), 14 pairs (15%) were discordant, and 6 pairs (6%) were concordant abnormal (Fig. 1). The discordant twin with normal amyloid-PET visual reads had higher BP_ND_ values compared to concordant twins with both normal amyloid-PET visual reads (p<0.05, Fig. 1H), suggesting that Aβ aggregation has already started in discordant normal twins, although still visually normal. Both amyloid abnormal and amyloid normal twins from discordant twin-pairs had higher BACE1 concentrations compared to concordant normal amyloid-PET twins (Fig. 1D), suggesting that increased BACE1 activity may indicate a very early event in sporadic AD^49^. The twin with normal amyloid from discordant pairs showed higher levels of t-tau and p-tau compared to concordant normal twin-pairs, which suggests that also tau levels are increased very early in the disease (Fig. 1I-J).

### CTCT and twin difference analyses

CTCT analyses showed that higher BACE1 and Aβ38 concentrations in one twin correlated with lower Aβ 1-42/1-40 ratios in the co-twin, and even stronger so with higher t-tau and p-tau levels (Aβ aggregation: r=-0.18--0.24, p=0.07-0.007; t-tau: r=0.56–0.58, p<0.0001; p-tau: r=0.32–0.54, p<0.0001-0.002, Table 4A). Within-pair differences in production markers were also related to within-pair differences in tau levels (t-tau: r=0.52-0.61, p<0.0001; p-tau: r=0.49-0.54, p<0.0001, Table 4B), but not to within-pair differences in Aβ aggregation markers. CTCT analyses and within-pair differences further showed that aggregation markers (i.e. lower Aβ 1-42/1-40 ratios and higher amyloid PET-BP_ND_ values) in one twin predicted higher t-tau and p-tau levels in their co-twin (Table 4).

**Table 4.** Cross-twin cross trait and within twin-pair difference analyses.

|  |  | A. Cross-twin cross-trait |  |  |  | B. Within twin-pair difference |  |  |  |
| --- | --- | --- | --- | --- | --- | --- | --- | --- | --- |
|  |  | Model 1 |  | Model 2 |  | Model 1 |  | Model 2 |  |
| | | R (SE);<br>95%CI | p-value | R (SE);<br>95%CI | p-value | $\beta$ (SE) | p-value | $\beta$ (SE) | p-value |
| **Association between amyloid aggregation markers** |  |  |  |  |  |  |  |  |  |
| Ratio CSF Amyloid-β 1-42/1-40 | PET global [18F]flutemetamol binding | -0.42 (0.08);<br>(-0.56 – -0.26) | <0.0001 | -0.33 (0.09);<br>(-0.49 – -0.15) | 0.0005 | -0.56 (0.13) | <0.0001 | -0.58 (0.14) | <0.0001 |
| **Associations between amyloid production and aggregation markers** |  |  |  |  |  |  |  |  |  |
| BACE-1 | Ratio CSF Amyloid-β 1-42/1-40 | -0.19 (0.10);<br>(-0.38 – 0.01) | 0.06 | -0.18 (0.10);<br>(-0.37 – 0.01) | 0.07 | -0.16 (0.14) | 0.26 | -0.13 (0.14) | 0.34 |
| Amyloid-β 1-38 | Ratio CSF Amyloid-β 1-42/1-40 | -0.36 (0.09);<br>(-0.53 – -0.16) | 0.0005 | -0.24 (0.10);<br>(-0.46 – -0.08) | 0.007 | -0.07 (0.14) | 0.61 | -0.05 (0.14) | 0.72 |
| Amyloid-β 1-38 | PET global [18F]flutemetamol binding | 0.23 (0.09);<br>(0.02 – 0.41) | 0.03 | 0.16 (0.10);<br>(-0.08 – 0.33) | 0.22 | 0.08 (0.16) | 0.63 | 0.08 (0.16) | 0.64 |
| **Associations between amyloid aggregation and tau markers** |  |  |  |  |  |  |  |  |  |
| CSF ratio Amyloid-β 1-42/1-40 | CSF t-tau | -0.53 (0.07);<br>(-0.66 – -0.37) | <0.0001 | -0.39 (0.09);<br>(-0.55 – -0.21) | <0.0001 | -0.40 (0.13) | 0.003 | -0.36 (0.13) | 0.009 |
| PET global [18F]flutemetamol binding | CSF t-tau | 0.37 (0.09);<br>(0.19 – 0.53) | 0.0001 | 0.25 (0.10);<br>(0.05 – 0.42) | 0.015 | 0.41 (0.13) | 0.002 | 0.40 (0.13) | 0.003 |
| CSF ratio Amyloid-β 1-42/1-40 | CSF p-tau | -0.54 (0.07);<br>(-0.67 – -0.37) | <0.0001 | -0.39 (0.09);<br>(-0.55 – -0.20) | 0.0001 | -0.53 (0.12) | <0.0001 | -0.53 (0.12) | <0.0001 |
| PET global [18F]flutemetamol binding | CSF p-tau | 0.36 (0.08);<br>(0.18 – 0.51) | 0.0001 | 0.22 (0.10);<br>(0.03 – 0.40) | 0.025 | 0.69 (0.10) | <0.0001 | 0.67 (0.10) | <0.0001 |
| **Associations between amyloid production and tau markers** |  |  |  |  |  |  |  |  |  |
| BACE-1 | CSF t-tau | 0.51 (0.08);<br>(0.35 – 0.65) | <0.0001 | 0.56 (0.07);<br>(0.41 – 0.69) | <0.0001 | 0.63 (0.11) | <0.0001 | 0.61 (0.11) | <0.0001 |
| Amyloid-β 1-38 | CSF t-tau | 0.65 (0.06);<br>(0.51 – 0.75) | <0.0001 | 0.58 (0.07);<br>(0.43 – 0.70) | <0.0001 | 0.53 (0.12) | <0.0001 | 0.52 (0.12) | <0.0001 |
| Amyloid-β 1-40 | CSF t-tau | 0.62 (0.07);<br>(0.47 – 0.73) | <0.0001 | 0.58 (0.07);<br>(0.42 – 0.70) | <0.0001 | 0.59 (0.11) | <0.0001 | 0.58 (0.11) | <0.0001 |
| BACE-1 | CSF p-tau | 0.48 (0.08);<br>(0.31 – 0.63) | <0.0001 | 0.54 (0.07);<br>(0.37 – 0.67) | <0.0001 | 0.50 (0.12) | <0.0001 | 0.49 (0.12) | <0.0001 |
| Amyloid-β 1-38 | CSF p-tau | 0.42 (0.09);<br>(0.23 – 0.58) | <0.0001 | 0.32 (0.10);<br>(0.12 – 0.50) | 0.002 | 0.48 (0.12) | <0.0001 | 0.49 (0.12) | <0.0001 |
| Amyloid-β 1-40 | CSF p-tau | 0.53 (0.08);<br>(0.37 – 0.67) | <0.0001 | 0.51 (0.08);<br>(0.35 – 0.65) | <0.0001 | 0.54 (0.12) | <0.0001 | 0.54 (0.12) | <0.0001 |
| **Associations between amyloid production markers** |  |  |  |  |  |  |  |  |  |
| BACE-1 | Amyloid-β 1-38 | 0.45 (0.09);<br>(0.26 – 0.60) | <0.0001 | 0.41 (0.09);<br>(0.24 – 0.58) | <0.0001 | 0.57 (0.11) | <0.0001 | 0.57 (0.12) | <0.0001 |
| BACE-1 | Amyloid-β 1-40 | 0.67 (0.06);<br>(0.54 – 0.77) | <0.0001 | 0.66 (0.06);<br>(0.53 – 0.77) | <0.0001 | 0.73 (0.09) | <0.0001 | 0.73 (0.10) | <0.0001 |
| Amyloid-β 1-38 | Amyloid-β 1-40 | 0.62 (0.07);<br>(0.48 – 0.73) | <0.0001 | 0.51 (0.08);<br>(0.41 – 0.69) | <0.0001 | 0.78 (0.09) | <0.0001 | 0.78 (0.09) | <0.0001 |
| **Associations between tau markers** |  |  |  |  |  |  |  |  |  |
| CSF t-tau | CSF p-tau | 0.68 (0.06);<br>(0.55 – 0.78) | <0.0001 | 0.57 (0.07);<br>(0.41 – 0.70) | <0.0001 | 0.74 (0.09) | <0.0001 | 0.74 (0.10) | <0.0001 |

CTCT = cross-twin cross-trait; CSF = cerebrospinal fluid; Amyloid- $\beta$ = amyloid beta; BACE = beta secretase; PET = positron emission tomography; t-tau = total-tau; p-tau = 181-phosphorylated-tau

### Comparison of CSF and PET Aβ markers, and t-tau with p-tau markers

We further studied the relationship between PET and CSF markers for Aβ aggregation, and observed moderately strong correlations between PET-BP_ND_ and the CSF Aβ 1-42/1-40 ratio across the total sample (β=-0.57 (SE=0.09), p<0.0001; Table 2), with CTCT (r=-0.33 (SE=0.09), p=0.0005; Table 4A) and within-pair difference analyses (β=-0.58 (SE=0.14), p<0.0001; Table 4B, Fig. 3). For t-tau and p-tau we observed high total sample correlations (β=0.85 (SE=0.08), which were attenuated in CTCT analyses r=0.57 (SE=0.07) and within-pair difference analyses (β=0.74 (SE=0.10), all p<0.0001, Tables 2, 4).

**Fig. 3.**
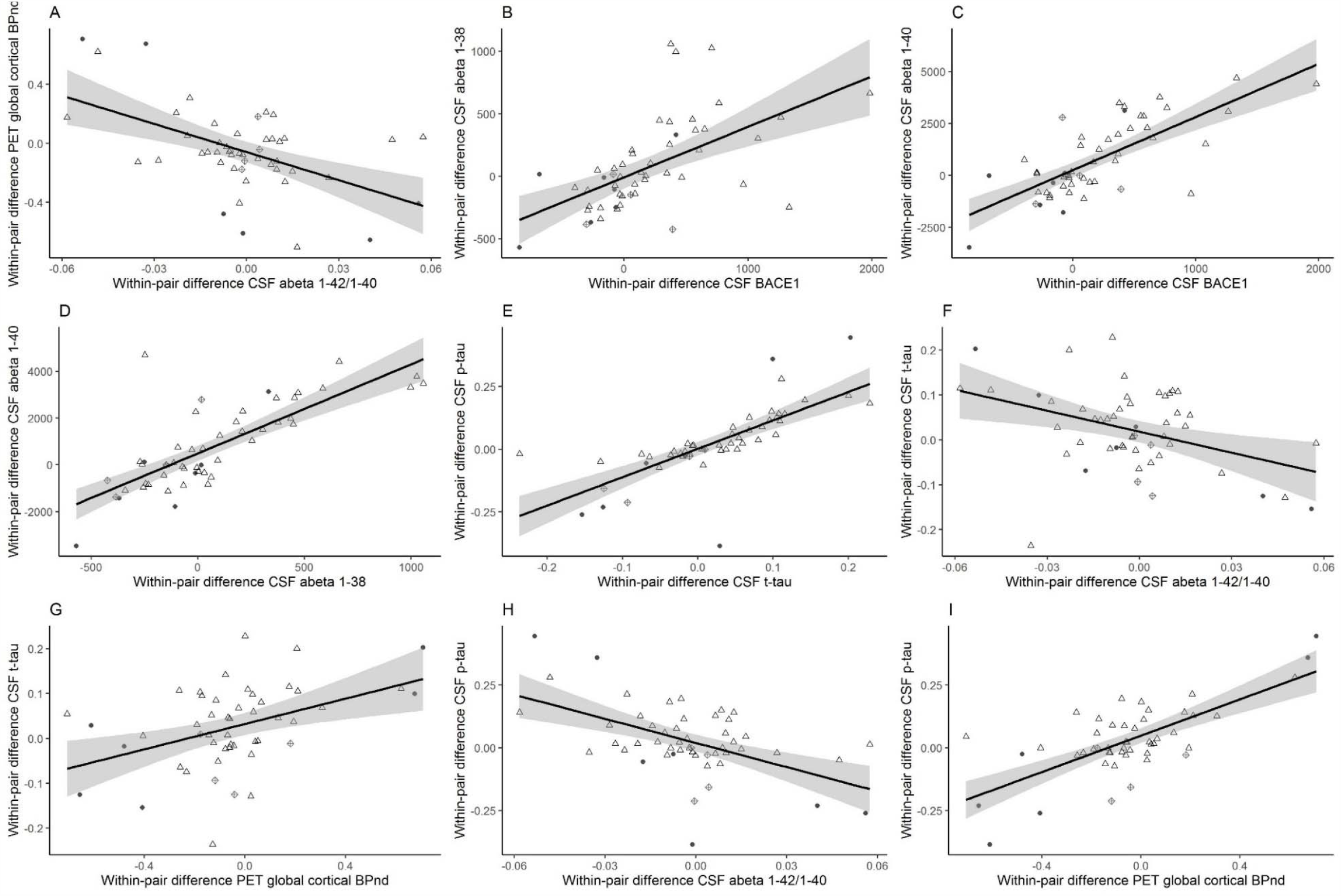
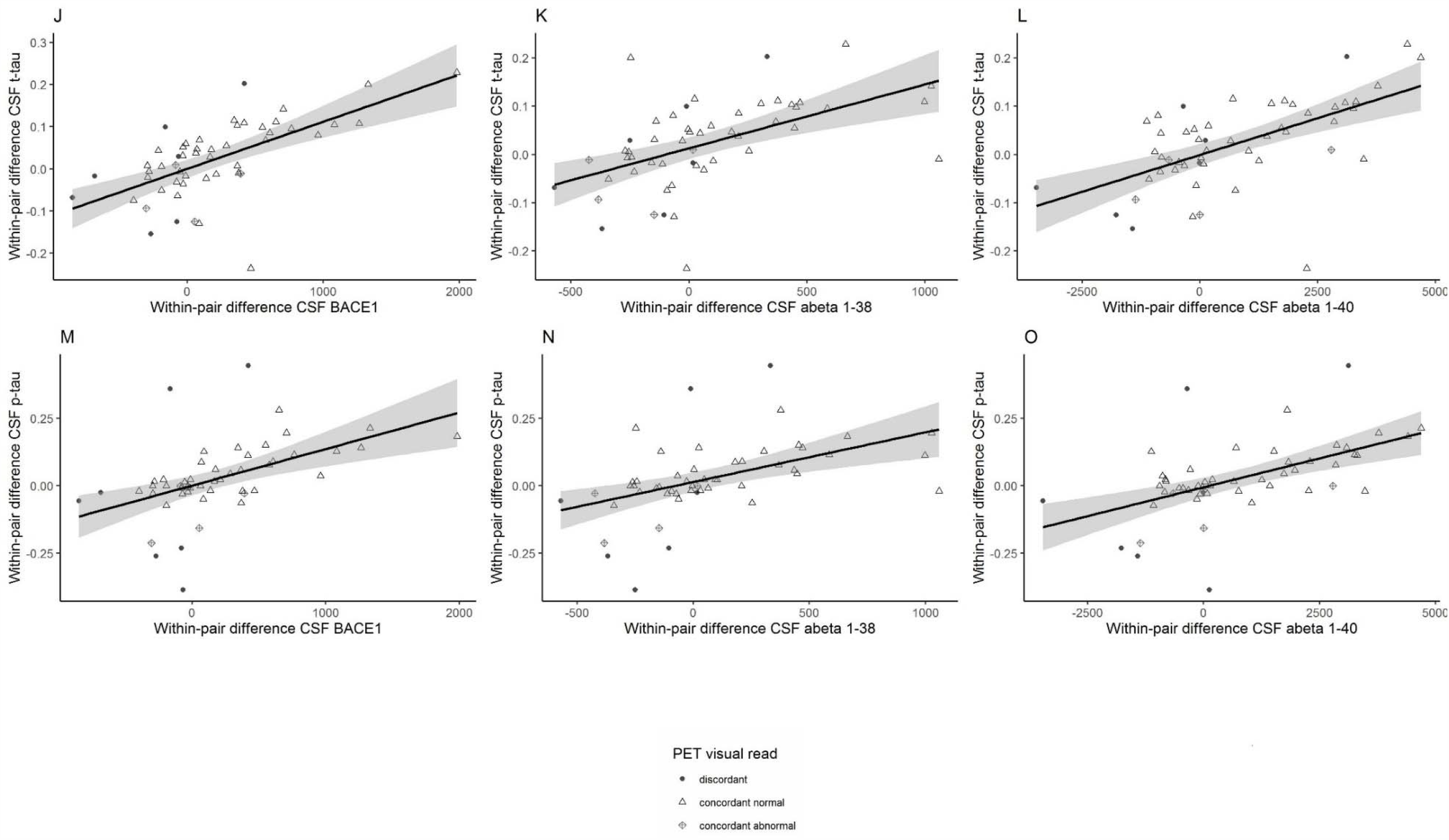
Monozygotic within-pair difference associations between amyloid aggregation markers, amyloid production markers and tau. Within-pair differences of (A) CSF amyloid-β 1-42/1-40 ratio with global cortical PET binding (BP_ND_); (B) CSF BACE1 with CSF amyloid-β 1-38; (C) CSF BACE1 with CSF amyloid-β 1-40; (D) CSF amyloid-β 1-38 with CSF amyloid-β 1-40; (E) CSF t-tau with CSF p-tau; (F) CSF amyloid-β 1-42/1-40 ratio with CSF t-tau; (G) global cortical PET binding (BP_ND_) with CSF t-tau; (H) CSF amyloid-β 1-42/1-40 ratio with CSF p-tau; (I) global cortical PET binding (BP_ND_) with CSF p-tau; (J) CSF BACE1 with CSF t-tau; (K) CSF amyloid-β 1-38 with CSF t-tau; (L) CSF amyloid-β 1-40 with CSF t-tau; (M) CSF BACE1 with CSF p-tau; (N) CSF amyloid-β 1-38 with CSF p-tau; (O) CSF amyloid-β 1-40 with CSF p-tau. Each dot represents one twin pair, twin pairs who are concordant normal on visual amyloid-PET read are shown as open triangles, twin pairs who are concordant abnormal on visual amyloid-PET read are shown as squares with a cross inside. Discordant pairs on visual amyloid-PET read are shown as black dots. Lower CSF ratio amyloid beta 1-42/1-40 and higher global cortical PET binding indicate more amyloid aggregation. CSF = cerebrospinal fluid; PET = positron emission tomography; BACE = beta secretase; t-tau = total-tau; p-tau = 181-phosphorylated-tau.

## 4. Discussion

Our study in cognitively normal older monozygotic twins suggests that in the very early stages of sporadic AD increased levels of Aβ production markers are associated with Aβ aggregation as well as higher levels of t-tau and p-tau. CTCT and twin difference analyses indicated that Aβ production and Aβ aggregation, Aβ production and tau markers, and tau and Aβ aggregation share, at least in part, a common underlying pathology. Furthermore, the moderately strong within twin-pair correlations for aggregation markers and the observation that 14 twin-pairs were discordant for amyloid pathology, suggests that environmental factors influence the start of Alzheimer’s disease pathological processes.

In ADAD it has been shown that mutations of the genes amyloid precursor protein (APP), presenilin 1 (PSEN1) or presenilin 2 (PSEN2) cause AD. These mutations can increase Aβ production, which leads to Aβ aggregation. To what extent increased Aβ production, decreased Aβ clearance, and other factors such as increased neuronal activity, increased inflammation or resistance to γ-secretase activity eventually leads to Aβ aggregation and neurodegeneration in sporadic AD remains largely unclear. Our findings suggest a role of increased Aβ production in the very early pathophysiology of sporadic AD. A particularly novel finding in our study is that Aβ production markers were correlated with t-tau and p-tau levels. Such relationships have previously been reported in studies using induced pluripotent stem cells (iPSC) models where the iPSC-derived purified neurons from sporadic AD showed significantly higher levels of Aβ40^50,51^, and tau^10^ compared to controls. The high CTCT and twin difference associations indicates that tau levels and Aβ production markers may share underlying pathophysiological processes. One such mechanism could be neuronal activity, which dynamically regulates APP cleavage and Aβ levels^52,53^. In mouse models, aberrant increased neuronal activity has been observed to precede plaque formation in AD^54^ and such hyperactivity has been related to increased neuronal secretion of both Aβ and tau, which compromises the metabolic homeostasis of neurons and contribute to network dysfunction^52,55^. We observed such changes in tau and amyloid towards abnormality very early in the discordant monozygotic twins who had still normal amyloid PET. We further found that Aβ production markers and tau levels have a shared genetic background. This warrants further investigation into genetic variants causing these changes in large combined genome-wide association studies with CSF.

We found different patterns in the discordant twin-pair analyses between Aβ production markers BACE1, Aβ40 and Aβ38. Possibly, this may indicate functional differences between secretases in production of Aβ peptides, since a previous study using ADAD iPSC-derived neurons showed that mutations of APP and PSEN1 have distinct effects on Aβ40 and Aβ38 levels by γ-secretase^56^. Future research should further investigate such effects in more detail, by measuring other markers that could reflect γ-secretase activity.

Furthermore, monozygotic twin-pair correlations for Aβ production markers and t-tau were much higher compared to those of aggregation markers and p-tau. Whether this high correlation resulted from shared genetics or shared environmental factors between twins could not be investigated as we did not include dizygotic twins in our study. However, previous studies indicated that shared environmental factors have a limited effect on neurological and neurodegenerative traits^4,57^. Future studies are needed to clarify the mechanisms that underlie the differences in amyloid production markers between cognitively normal individuals as this may provide novel clues for reducing Aβ aggregation and tau phosphorylation.

Monozygotic twin-pair correlations observed for CSF and PET Aβ aggregation markers and p-tau ranged between 0.49 and 0.52, which similar to previous CSF and PET concordance estimates reported by previous, smaller studies in cognitively normal individuals^6,7^, and lower compared to twin similarity estimates for AD-type dementia (up to 79%)^4^. It is possible that monozygotic twin similarities in Aβ aggregation may be lower because of the early disease stage, as the similarity between twins for AD-type dementia increases with age^4^. As such, it is likely that in amyloid discordant pairs, the twin with a normal amyloid-PET visual read may become abnormal in the future, and the notion that such individuals already showed higher BP_ND_ values compared to twin-pairs with both normal amyloid-PET supports this explanation. CTCT and twin difference associations indicated that CSF and PET measures of amyloid aggregation reflect a common underlying pathophysiology. However, associations were moderate, in line with earlier studies^58^, suggesting that they may also capture different aspects of aggregation.

Bigger sample sizes for investigating very early pathophysiology of Alzheimer’s disease that include more individuals with abnormal amyloid would be desirable to increase statistical power. However, our older monozygotic twin approach is unique, and has enabled us to estimate the contribution of genetic and environmental factors to the start of Aβ aggregation. Longitudinal research is needed to investigate whether twin-pairs discordant for amyloid abnormality will become more similar in their levels of Aβ aggregation and p-tau over time^4^. Furthermore, aberrant CSF Aβ1-42/1-40 ratios are evident for AD pathology, but it still remains unclear what differences in CSF Aβ production markers represent in sporadic AD. In conclusion, previous population studies have indicated that environmental factors influence dementia risk^59^ and our results show that this, at least partially, may be explained through the impact on Aβ and tau aggregation. Identification of such environmental factors may lead to new ways to prevent AD. In addition, identification of genes and mechanisms associated with high amyloid production in cognitively normal individuals may further provide novel leads to reduce Aβ aggregation.

## Data Availability

The data that support the findings of this study are available from the corresponding author after signing a material transfer agreement.

## Acknowledgments

This work has received support from the EU/EFPIA Innovative Medicines Initiative Joint Undertaking (EMIF grant n° 115372). This work received in kind sponsoring of the CSF assay from ADx NeuroSciences and Euroimmun, and the PET-tracer [^18^F]flutemetamol from GE Healthcare. We thank all participating twins for their dedication.

## Author contributions

EK, JT, MtK, and AdB, collected data. EK, MtK and MMY performed image analysis and SDM and CET CSF analysis. EK, JT, AdB and MGN performed statistical analyses. EK, JT, AdB, BMT and PJV drafted the manuscript. BMT, MtK, MMY, SDM, MGN, HV, AAL, CET, BNMvB, DIB, PHS, and PJV edited the manuscript for critical content. PJV conceived the study, designed the protocol, and provided overall study supervision. All authors read and approved the final version of the manuscript. HV is a co-founder of ADx NeuroSciences and a founder of Biomarkable bvba.

## Conflicts of interests

The authors declare the following competing interests:

HV is working on behalf of Biomarkable bvba for ADx NeuroSciences, who provided the ELISA’s used in this study.

The other authors do not have a competing interest.

